# The tumor burden of metastatic colorectal cancer patients at initial diagnosis, pre- versus post-Covid-19 lockdown

**DOI:** 10.1101/2021.03.17.21253408

**Authors:** AR Thierry, B Pastor, E Pisareva, F Ghiringhelli, O Bouche, C De La Fouchardière, J Vanbockstael, D Smith, E François, M Dos Santos, D Botsen, S Ellis, M Fonck, T Andre, E Guardiola, F Khemissa, B Linot, J Martin-Babau, Y Rinaldi, E Assenat, L Clavel, S Dominguez, C Gavoille, D Sefrioui, V Pezzella, C Mollevi, M Ychou, T Mazard

## Abstract

**Background:** The COVID-19 pandemic led to a significant reduction in the provision of screening, case identification and hospital referrals to cancer patients. To our knowledge, no study has yet correlated quantitatively the consequences of these limitations for cancer patient management. This study evaluates the implications of such reductions for patients newly diagnosed with metastatic colorectal cancer (mCRC) in both the pre- and post-lockdown periods.

**Methods:** We examined 80 newly identified mCRC patients from 18 different clinical centers. These cases come from the screening procedure of a clinical trial which is using circulating DNA (cirDNA) analysis to determine their *RAS* and *BRAF* status.

**Results:** The tumor burden as evaluated by the median total plasma cirDNA concentration showed a statistically higher level in patients diagnosed post-lockdown compared to those diagnosed pre-lockdown (119.2 versus 17.3 ng/mL; p<0.0001). In order to link tumor burden to survival, we compared the survival of these mCRC patients with previous studies in which cirDNA was examined in the same way (median survival, 16.2 months; median follow up, 48.7 months, N=135). Given the poor survival rate of mCRC patients with high cirDNA levels (14.7 vs 20.0 and 8.8 vs 19.3 months median survival when dichotomizing the cohort by the median cirDNA concentration 24.4 and 100 ng/mL, respectively), our study points to the potential deleterious consequences of the COVID-19 pandemic.

**Conclusions:** Recognizing that our exploratory study offers a snapshot of an evolving situation, our observations nonetheless clearly highlight the need to determine actions which would minimize delays in diagnosis during the ongoing and future waves of COVID-19.

## Introduction

The unprecedented burden placed on health systems worldwide by the coronavirus disease 2019 (COVID-19) crisis has had numerous and significant implications for cancer care^1,2,2^. People have been more reluctant to come to health services for fear of infection, particularly patients already diagnosed with cancer, given that cancer is considered a comorbidity. Cuts to (or indeed the suspension of) screening programs and diagnostic services in many countries have caused delays in diagnosis^1,3,4,5^. To reduce the risk of SARS-Cov-2 exposure for cancer patients during therapy procedures, access to treatment has been restricted^3^. Lastly, the reprioritization of human resources and equipment to COVID-19 pandemic management has also resulted in the provision of sub-optimal or delayed care^6,1^.

These implications have been exacerbated by the COVID containment measures enacted by different countries, which have tended to evolve from local and national recommendations and restrictions to local and national lockdowns^1,2^. Such measures were initially seen in the first months of 2020 in Asia and Oceania, followed by Europe and the Americas in March, depending mainly on the date of the first SARS-Cov-2 infection cases in those areas^2^. The number of COVID patients necessitating hospitalization and critical care has placed an unprecedented burden on health services, and consequently limited their resources. Individual fears of contracting the virus, coupled with restrictions imposed on movement by the authorities, have generated additional barriers - both physical and psychological - for patients who need to access essential care.

To explore the indirect health effects of the COVID-19 pandemic on newly diagnosed cancer patients, we set out an observational study. To our knowledge, no other clinical evaluation exists of the impact of COVID-19 lockdowns on the tumor burden of cancer patients at initial diagnosis. We used cirDNA analysis to assess the patients’ tumor burden. CirDNA is newly-identified source of biological information, which has attracted the attention of researchers and clinicians in numerous fields^7^. Its clinical potential in oncology is significant, and includes molecular profiling, detection of residual disease, control of treatment efficacy, detection of clonal resistance, surveillance of recurrence, and screening^8^. CirDNA first showed its promise in contributing to companion tests as a “liquid biopsy;” it then acquired European Medicines Agency (EMA) approval for use in the detection of sensitizing and/or resistant somatic alterations in oncodrivers, such as in lung cancer and melanoma, as a tool to guide clinicians in selecting targeted therapies^9,8^. Numerous works report that tumors secrete DNA into the blood stream in quantities proportional to their masses^10,11,12^ especially in the case of metastatic colorectal cancer (mCRC)^10,13,14,15^. Thus, cirDNA offers analytical and clinical advantages over conventional antigenic biomarkers such as CEA, and may be considered as a surrogate marker of disease progression, at least in mCRC^16,13,17,14,18,19^ (Suppl. Fig. 1).

## Methods

In this study, we included patients from the screening procedure of the ongoing UCGI 28 PANIRINOX study (NCT02980510/EudraCT n°2016-001490-33), who were recruited before and after the lockdown enacted in France in the spring of 2020. The PANIRINOX study is a first line randomized phase II study, assessing the activity of a combination chemotherapy with fluorouracil, leucovorin, oxaliplatin and panitumumab ± irinotecan (FOLFOX + panitumumab vs FOLFIRINOX + panitumumab) in unresectable mCRC patients, selected by *RAS* and *B-RAF* status obtained from cirDNA analysis. It is the first interventional study to use cirDNA as a companion test for selecting mCRC patients towards anti-EGFR targeted therapy (Suppl. Information). That trial involves 31 French hospitals and cancer centers. The primary endpoint is the complete response rate, defined as the complete disappearance of metastatic lesions and CEA level normalization after a maximum of 12 treatment cycles Other major selection criteria are: age (between 18 and 75 years); ECOG performance status 0 or 1; absence of previous treatment for metastatic disease; and absence of use of oxaliplatin in an adjuvant setting (Suppl. Information). After they give written informed consent, patients are screened through a blood-sampling procedure to determine their *RAS* and *BRAF* tumor status according to plasma analysis of circulating cell free DNA, using Intplex technology^20,21^. Tumors considered as *RAS*/*BRAF* wild type are subsequently included in the PANIRINOX study if they fulfill all other inclusion criteria (Suppl. Information).

That ancillary study, therefore, benefits from the accuracy with which cirDNA can evaluate tumor burden, and from the trial’s rigorous inclusion procedure and reporting, all of which supports the accuracy of assessment needed to achieve our objective. We examined all screened pre- and post-lockdown patients irrespective of their *RAS* and *BRAF* mutational status, in order to preclude any potential bias linked to mutational status. Given the interventional impact of cirDNA analysis in the PANIRINOX study, it is completed within five days of reception of the blood samples. In addition to cirDNA parameters analysis, we also collected, at the same time, demographic and clinico-biological parameters known to have prognosis value in this setting^22,23^.

## Results

In France, the first mandatory home lockdown of 2020 lasted 55 days, from 17 March to 11 May. The PANIRINOX study screening was consequently interrupted on 19 March and resumed on 11 May (a 53-day interruption). Our study compared the cirDNA values determined in all patients (N=40) screened from the May 14, 2020 up to September 3, 2020 (a 110 day period) to those found in all patients (N=40) screened before the lockdown (from 11 November 2019 up to 9 March 2020). Pre-analytical conditions of the cirDNA analysis followed strict guidelines and methodologies which have been previously validated^24,20,21,25^. As observed in Figure 1, the cirDNA concentration median is 17.3 and 119.2 ng/mL, before and after the lockdown, respectively (Suppl. Table 1). This represents a nearly 7-fold increase. There is a high statistical difference between the two cohorts (P<0.0001) (Figure 1 and 2). The values obtained in the patients included before lockdown (N=40) are similar to those obtained in all patients previously included from the start of the PANIRINOX study (N=188; starting 30 months before lockdown), showing a median cirDNA concentration of 13.0 ng/mL plasma (Figure 2). In addition, the cirDNA concentration values determined in all the 110 days fractional cohorts before lockdown from the start of the PANIRINOX study showed no statistical difference with the pre-lockdown study cohort (N=40) whereas they are statistically different as compared to the post-lockdown study cohort (N=40) (Figure 2).

**Figure 1:**
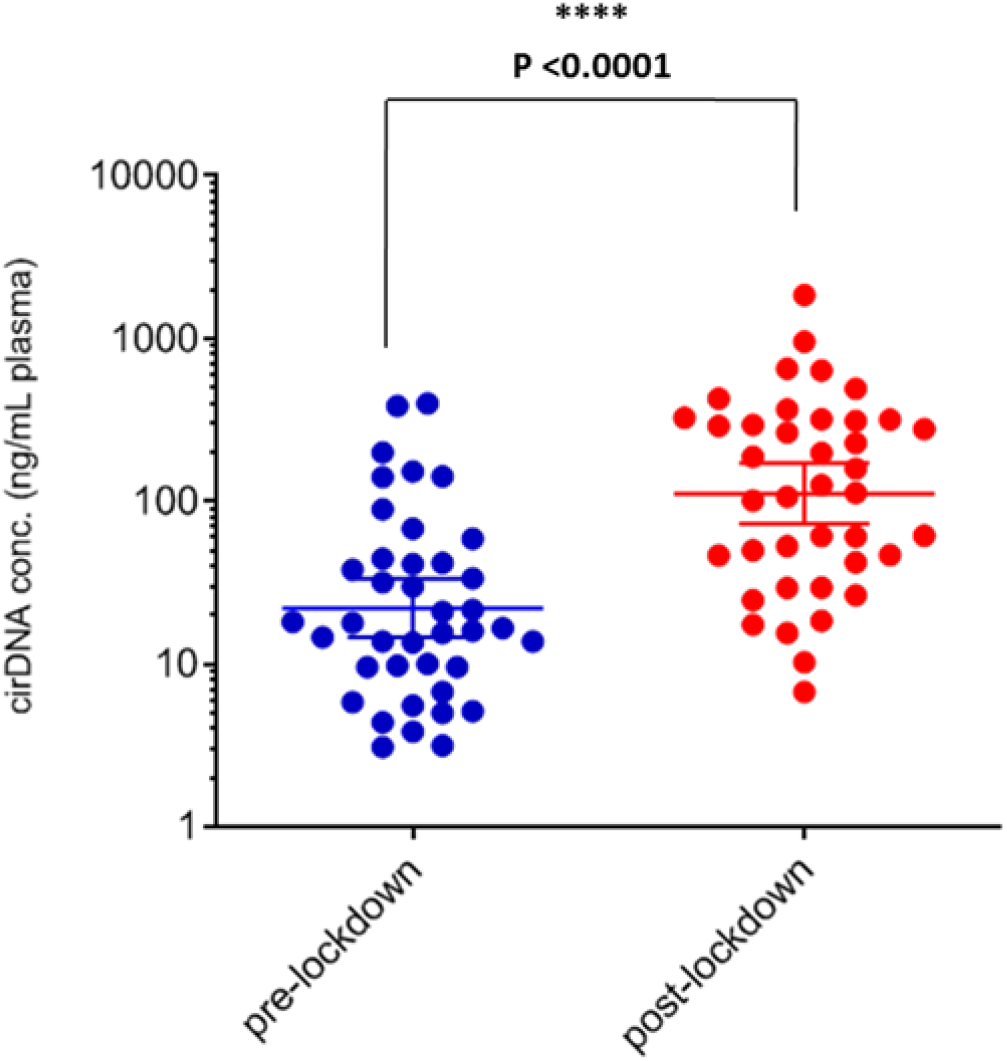
Comparison of cirDNA concentration in newly diagnosed mCRC patients from the pre- and post-lockdown study cohorts. Group median is represented by a horizontal bold bar. Mann-Whitney U test was performed to compare these distributions, and revealed a significant difference between pre-and post-lockdown patients (with p-value<0.0001). Each dot (blue, pre-lockdown; red, post-lockdown) represents the values of a single patient.

**Figure 2:**
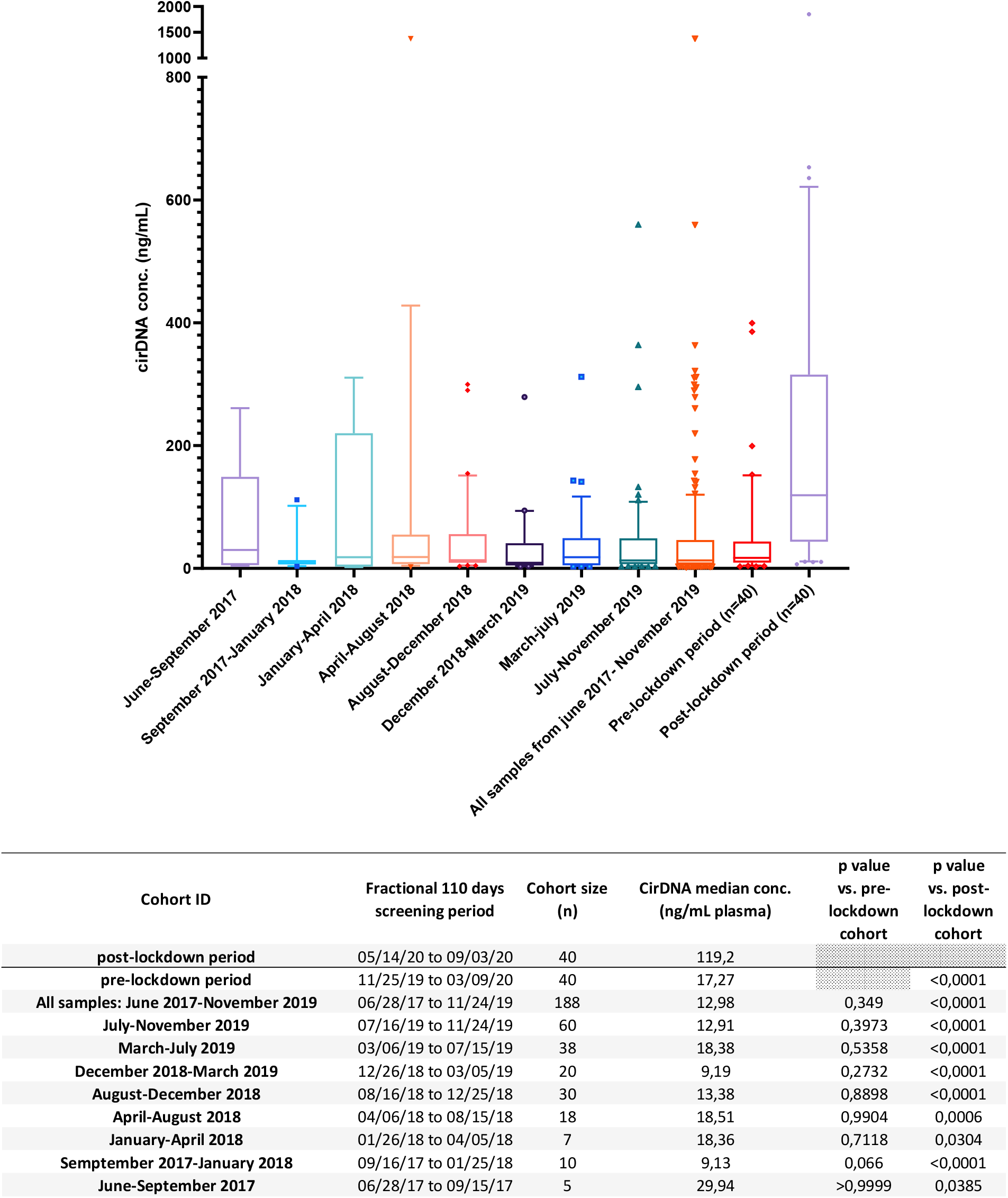
Comparison of study cohort of fractional 110 day period groups of patients initially screened from the start of the PANIRINOX study with the pre-lockdown study cohort. Top, box plot representation of cirDNA concentration values according to fractional 110 day period groups, pre- and post-lockdown study cohorts. Mann-Whitney U test was performed to compare distribution. Bottom, description of the fractional 110 day period groups of patients, and statistical characterization of the difference of median cirDNA concentration as determined in the post-lockdown study cohort and the fractional 110 day period groups of patients.

This increase seen in cirDNA values post-lockdown is striking, and points to levels of tumor burden at diagnosis which have been shown to affect patient survival ^26,27,28^. To estimate the potential impact on survival of diagnostic delays arising from to lockdown, we retrospectively analyzed data accumulated during our last two clinical studies^20,21^ from which we identified “all comers” newly diagnosed mCRC patients, and in which a rigorously identical methodology was used to assess cirDNA (N=135) before patients began first line chemotherapy. The median cirDNA concentration (24.4 ng/mL, range [2.3-1406], Suppl. Table 2) of these patients is similar to that observed in the pre-lockdown cohort studied from the PANIRINOX trial. As observed in Figure 3A, mCRC patient median survival in those two clinical studies (performed after a median follow up of 48.7 months, 95% CI [43.3-55.4]) is 16.2 months (N=135; 95% CI [13.6-20.6]). The median survival rate is low when compared with that seen in the latest literature up-to-date literature. This may be attributed to the starting date of these two clinical studies (2009 and 2014), and to a less stringent patient selection compared to the PANIRINOX study; it nonetheless fits within the median survival range as previously observed in a meta-analysis^29^ (16-23 months). When dichotomizing this cohort by the cirDNA concentration median value (24.4 ng/mL, range [2.3-1406]), patients diagnosed with higher cirDNA plasma amounts showed a statistically lower median survival (14.7 (95% CI [8.8-18.0]) vs 20.0 (95% CI [14.1-32.0]) months, HR: 1.74, 95% CI [1.2-2.6]; P=0.005) (Figure 3B, Suppl. Table 2). When dichotomizing the cohort by 100 ng/mL, patients diagnosed with higher cirDNA plasma amounts also showed a statistically lower median survival (8.80, 95% CI [2.8-14.0] vs 19.3, 95%CI [2.8-14.0]) months; HR: 2.00, 95% CI [1.2-3.3] P=0.009) (Figure 3C, Suppl. Table 2). The comparative study of the lockdown impact (Figure 1) revealed 23 out of 40 patients (58%) showing a cirDNA amount over 100 ng/mL post-lockdown, and only 6 out of 40 patients (15%) above this concentration in pre-lockdown period. This could mean that 17 (23 minus 6) or 43% (58% minus 15%) supplementary patients diagnosed post-lockdown would have a median survival which is 54% (from 19.3 to 8.8 ng/mL) less than that of patients diagnosed pre-lockdown, presuming care management was equivalent, despite the difference in time period between survival cohort and study cohort. We are aware that this remains an assumption, but one which nonetheless illustrates and anticipates the lockdowns’ marked deleterious impact on these patients’ health. The full lockdown-related impact on patient survival will of course be determined over a three year survival study.

**Figure 3:**
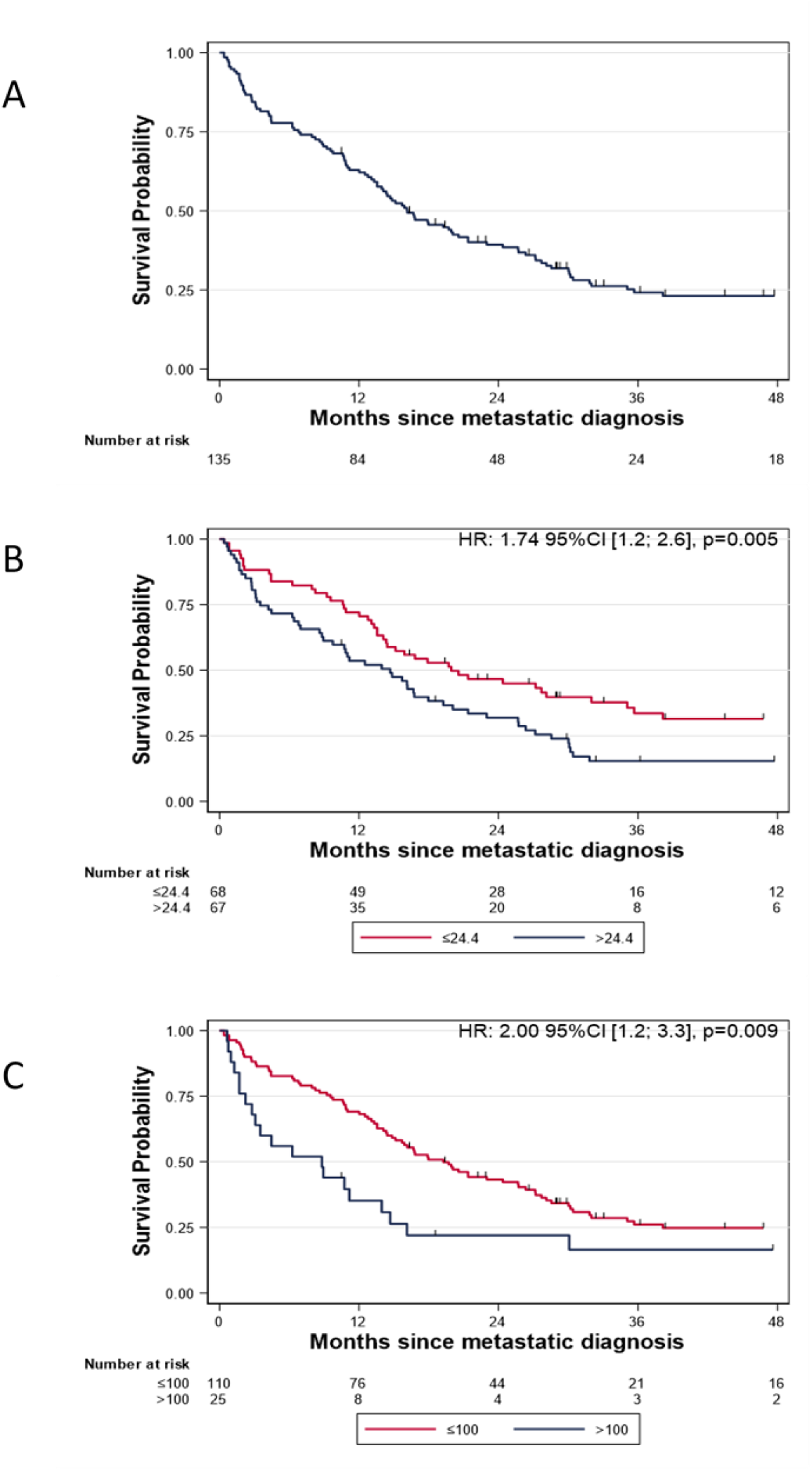
Overall survival analysis on the retrospective cohort of newly diagnosed mCRC patients. A, Kaplan-Meier survival curve on the full cohort (N=135). B, Kaplan-Meier survival curve and log-rank test according to cirDNA concentrations dichotomized around the median (24.4 ng/mL). C, Kaplan-Meier survival curve and log-rank test according to cirDNA concentrations dichotomized around the median (100 ng/mL).

Regarding patient characteristics, no difference was seen in the groups screened pre- and post-lockdown (Table 1, Suppl. Figure 2-3). The delay of blood sample delivery was also similar, as was the mutant cirDNA concentration, and the mutant allele frequency (Suppl. Figure 4-6, Suppl. Table 1). The white blood cell count, LDH, and CEA median level were slightly higher in the post-lockdown setting, but without the difference being statistically significant (Table 1, Suppl. Figure 7-9). Note, the cirDNA concentration significantly correlates with the LDH and white blood cell count (r=0.72, p<0.0001; and r=0.73, p<0.0001, respectively) in the post-lockdown patients, while the CEA does not (Suppl. Figure 10-14).

**Table 1.**
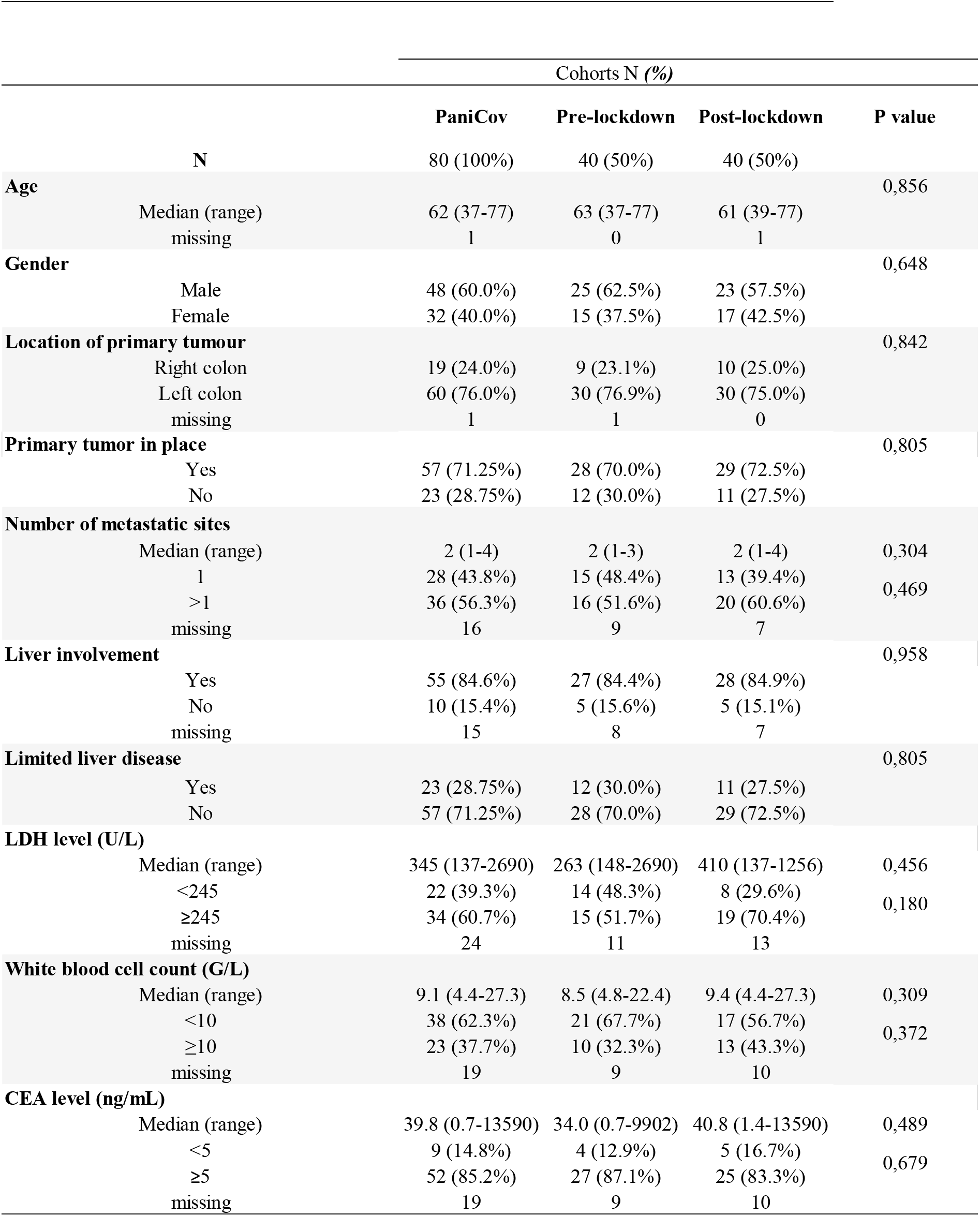
Patients characteristics.

## Discussion

In light of the differential in tumor burden between patients diagnosed pre- and post-lockdown, and the resulting risk of reduced survival, our data points to the deleterious impact of the lockdown and pandemic-related circumstances on newly diagnosed mCRC patients who delayed their first visit to an oncologist.

The lower number of diagnoses performed during the COVID-19 crisis^1,3^ is undoubtedly due to patients’ reluctance to visit a doctor, arising from fears of catching COVID-19 or of burdening the heath system. Take the example of a Australian sixty year-old patient, currently being treated for jaw cancer, who said: “*Because I didn’t think it was an emergency, I didn’t make any moves to go back to see anybody. And also because it was in the middle of COVID I was afraid of getting COVID or giving COVID, and so I just put up with it for a while*.”^30^. In addition to patients’ subjective anxieties and reticence, there have been numerous reports of significant delays in sending out of millions of solicitations for bowel cancer screening, and of a backlog (in England alone) of thousands of individuals waiting for further investigation after receiving a positive screening result^3,2^.

While the lockdown was undoubtedly a necessity, it led to unintended consequences in the diagnosis of various cancers. While the pandemic has dramatically affected all aspects of the cancer care pathway those especially affected are screening, diagnosis and surgical treatment^31,32,4^. For instance, De Vincentiis et al have reported that the number of cancer diagnoses in Italy decreased by 39% in the first semester of 2020, compared to the average number recorded in 2018 and 2019^4^. The highest declines in diagnosis were observed in prostate cancer (75%), bladder cancer (66%), and CRC (62%), as determined by comparing the number of new or first metastatic malignant diagnoses during the lockdown (weeks 11–20 of 2020) to the same period in the previous two years. Given that colonoscopy numbers are closely related to initial CRC diagnosis, it is interesting to note that there was a 55% drop in colon exams between March and April of 2020, as reported by “Cancer Australia.” In addition to the abrupt drop (86%) of preventive CRC screenings in the United States following the declaration of the COVID-19 national emergency (March 1, 2020), a drop of 64% (i.e. 95,000) in the number of colonoscopies performed between March 15 and June 16, 2020, as compared with previous years, has recently been reported^33^. Furthermore, after June 16, weekly volumes remained 36% lower than pre-COVID-19 levels^33^. With particular relevance to our study, it should be noted that an observational Taiwanese cancer registry study^34^, based on 39,000 newly identified CRC cases, reports that the risk of death increases significantly relative to the delay between diagnosis and treatment: to an interval ≤ 30 days, for 31–150 days: HR 1.51; (95% CI 1.43 to 1.59); for ≥151 days: HR 1.64; (95% CI 1.54 to 1.76). The French ONCOCARE-COV study confirmed a reduction in terms of CRC FIT screening (−86%), CRC biomolecular somatic analyses (−59%), and new patients files discussed in multidisciplinary tumor board meeting (−39%) during the 3-month lockdown period in 2020 compared with the same trimester in 2019^31^.

Several papers have generated model-based estimates of the clinical consequences of delaying the first visit of newly diagnosed cancer patients^26,35,36^. In the UK, an even modest delay of three to six months in surgery for cancer is expected to mitigate 19%-43% of the life years gained by hospitalization, as reported by Sud et al^35^. Recent estimates from Lai et al^37^predict about 18,000 excess cancer deaths over the next 12 months due to the COVID-19 crisis. Besides the 1 million deaths from breast cancer alone which would have been expected over the next decade in the US, about 10,000 additional deaths are now expected due to pandemic-related delays of no more than six months in screening and cancer care^38^.

The COVID-19 related lockdowns’ effects on the provision of health care were particularly notable in oncology, and the prospect of repeated or extended lockdowns entails the threat of decreased surveillance and advance care planning. To address this, recommendations and guidance for delivering care to cancer patients during the crisis and lockdowns were established by regulatory institutions such as the ASCO or ESMO^1,2^. In order to minimize risks to patients with gastrointestinal malignancies, for instance, new priorities were set by the American College of Surgeons, the Society of Surgical Oncology, the French digestive oncology intergroup guideline (TNCD), and the ESMO; these included prioritizing surgery for colon cancer involving imminent obstruction, or for locally advanced rectal cancer; similarly, new priorities concerning CRC management were set by the US Colorectal Cancer Alliance, TNCD, NCCN, ESMO, and the City of Hope National Medical Center^1^. Such recommendations were used to reclassify and reprioritize ongoing CRC care and management during the lockdown. When CRC is diagnosed early, the prognosis for treatment and cure is more favorable. On the basis of a large meta-analysis, Hanna et al.^26^ recently reported that even a four week delay of treatment is associated with increased mortality for seven cancers, in particular CRC (HR, 1.04; 95% CI [0.95-1.13]). Our quantitative observation (albeit performed on a small sample of a specific type of cancer patient) shows that delays in diagnosis would unnecessarily cost lives and life-years. At a time when France and several other European countries are enacting a second lockdown in response to a new proliferation of the virus, we believe that corrective action to the impact on cancer diagnosis should be taken, including (i), reinforcing mass screening using the faecal occult blood test; (ii), increasing the communication strategy needed to avoid late patient diagnosis; and (iii), the provision of adequate resources and robust plans to deal with backlogs in diagnosis and treatment. We speculate that patient triage could be performed based on the quick assessment of tumor burden and the testing of biomarkers with predictive and prognosis value (such as immunohistochemistry for mismatch repair proteins; mutation analysis for *KRAS, NRAS, and BRAF)*. For this purpose, we believe that cirDNA analysis revealing qualitative (tumor molecular profiling) or quantitative information^8,20 21,39,13^ may represent an ideal tool, as previously reported^16,19,40^.

Despite the growing number of reports which highlight the magnitude of the indirect effects of the pandemic on health systems worldwide, no direct evaluation has yet been made regarding the increased tumor burden of newly diagnosed patients post-lockdown. Ours is the first assessment of the impact on diagnostic services related to a specific cancer. It would be premature to envisage a definitive evaluation of the fallout from all delays in diagnosis, screening and treatment, and our exploratory study is offered rather as a snapshot of a situation which continues to evolve. In conclusion, our observation at least point out CRC as a major area for intervention in order to minimize the clinical impact of a diagnostic delay due to COVID-19 crisis.

## Data Availability

The data are freely available

## Acknowledgements

The authors thank the excellent technical assistance of C. Sanchez and A. Kudriatsev, and Cormac Mc Carthy for English editing. We thank also all CRA and clinical co-investigators, M. Bergeaud (Unicancer, Paris), and the support of AMGEN. B. Pastor was partially supported by SIRIC Montpellier Cancer Grant INCa_Inserm_DGOS_12553, and AR Thierry by INSERM.

## Supplementary Information

## Materials and methods

### 1. PANIRINOX general information

Groupe UNICANCER Tumor Group: UCGI; Protocol n°: UC-0110/1608; EudraCT n°: 2016-001490-33 INDICATION: RAS and B-RAF wild-type metastatic colorectal cancer. *RAS* and *B-RAF* mutation will be determined using cirDNA by IntPlex method.

METHODOLOGY: National trial, multicenter, randomized, phase II assessing combination chemotherapy with fluorouracil, leucovorin, oxaliplatin and panitumumab ± irinotecan (FOLFOX + panitumumab vs FOLFIRINOX + panitumumab) in metastatic colorectal cancer patients selected by *RAS* and *B-RAF* status from cirDNA analysis.

PRIMARY OBJECTIVE: Evaluation of complete response rate of treatment combining FOLFIRINOX and panitumumab.

SECONDARY OBJECTIVE(S):

- Overall Survival
- Progression free survival
- Secondary resection
- Early tumor shrinkage (ETS)
- Depth of response (DpR)
- Safety profile (NCI CTCAE v 4.03 classification)
- Diagnostic performance of ccfDNA analysis compared to the tumor-tissue analysis (current gold standard).

INCLUSION CRITERIA:

1. Age between 18 and 75 years
2. ECOG PS between 0 and 1
3. Histologically confirmed adenocarcinoma of the colon or rectum
4. Untreated synchronous or metachronous metastatic disease deemed unresectable with curative intent
5. K-Ras (codons 12, 13, 59, 61, 117, 146), N-Ras (codons 12, 13, 59, 61) and B-Raf (codon 600) wild-type tumor status according to plasma analysis of circulating cell free DNA by Intplex technology
6. Measurable disease according to RECIST version 1.1
7. Adequate hematologic, hepatic and renal functions:
  - Absolute neutrophil count (ANC) ≥2 x 109/L
  - Haemoglobin ≥9 g/dL
  - Platelets (PTL) ≥100 x 109/L
  - AST/ALT ≤5 x ULN
  - Alkaline phosphatase ≤2.5 x ULN
  - Bilirubin ≤1.5 x ULN
  - Creatinine clearance ≥50 mL/min (Cockcroft and Gault formula)
8. Life expectancy of at least 3 months
9. Adequate contraception if applicable
10. Patient affiliated to a social security regimen
11. Patient information and signed written consent form
12. Uracilemia < 16 ng/ml

NON INCLUSION CRITERIA:

1. History of other malignancy within the previous 5 years (except for appropriately treated insitu cervix carcinoma and non-melanoma skin carcinoma)
2. Adjuvant treatment with oxaliplatin
3. Previous treatment for metastatic disease
4. Patients who received any chemo- and/or radiotherapy within 15 days from the date of blood sampling for the RAS and BRAF test
5. Brain metastases
6. Patients with a history of severe or life-threatening hypersensitivity to the active substances or to any of the excipients delivered in this study
7. Patients with history of pulmonary fibrosis or interstitial pneumonitis
8. Previous organ transplantation, HIV or other immunodeficiency syndromes
9. Concomitant medications/comorbidities that may prevent the patient from receiving study treatment as uncontrolled intercurrent illness (for instance: active infection, active inflammatory disorders, inflammatory bowel disease, intestinal obstruction, symptomatic congestive heart failure, uncontrolled hypertension…)
10. Persistent peripheral neuropathy >grade1 (NCI CT v4.03)
11. Ionic disorders as:
  - Kalemia ≤1 x LLN
  - Magnesemia <0.5mmol/L
  - Calcemia <2mmol/L
12. Patient with known dihydropyrimidine dehydrogenase deficiency
13. QT/QTc>450msec for men and >470msec for women
14. Patient with contraindication for trial drugs
15. Concomitant intake of St. John’s wort
16. Other concomitant cancer
17. Participation in another therapeutic trial
18. Pregnant woman or lactating woman
19. Patients with psychological, familial, sociological or geographical condition hampering compliance with the study protocol and follow-up schedule
20. Legal incapacity or limited legal capacity

## 2. CirDNA analysis

All blood samples were collected in 10-milliter (mL) Streck tubes in all 27 clinical centers that sent by express mail the day or the day after blood draw following strict procedural guidelines. The IRCM U1094/ INSERM laboratory received blood samples within 5 days post-blood draw and centralized all following procedures towards cirDNA analysis under stringent pre-analytical guidelines reported by the laboratory^24^. The blood was then centrifuged at 1,200 g at 4°C for 10 minutes. The supernatants were isolated in sterile 1.5 mL Eppendorf tubes and centrifuged at 16,000 g at 4°C for 10 minutes. Afterwards, the plasma was either immediately used for DNA extraction or stored at -20°C. CirDNA was extracted from 1 mL of plasma using the QIAmp DNA Mini Blood kit (Qiagen) according to the ‘‘Blood and body fluid protocol.’’ DNA extracts were kept at -20°C until used^25^.

The PANIRINOX clinical study implies the use of the IntPlex methodology to select patients showing WT mutational status for *KRAS, NRAS* and *BRAF* genes. IntPlex was established by our team and is based on an allele-specific blocker Q-PCR–based method specific for cirDNA analysis to enable the detection of point mutation and to determine mutant allele concentration^20^. This method combined the use of (1) allele-specific Q-PCR with blocking 3′-phosphate–modified oligonucleotide; (2) low Tm primers with mutation in 3′; (3) an integrated primer design; (4) routine internal positive and negative controls, and (5) optimal analytical procedures. This method enables the determination of the presence of a mutation, as well as the total concentration of cirDNA, the concentration of mutant cirDNA, the mutant allele frequency, and an index of DNA integrity. Note, the total cirDNA concentration value is determined by targeting a *KRAS* WT sequence, and is internally controlled by targeting a *BRAF* WT sequence for each sample^25^.

CirDNA quantification methodology and the data description were carried out according to the MIQE guidelines^20^. Q-PCR amplifications were carried out at least in triplicate in a 25-μl reaction volume on CFX96 thermocycler (Bio-Rad) using the Bio-Rad CFX manager. Each PCR mixture was composed of 12.5 μl of PCR mix (Bio-Rad Supermix SYBR Green), 2.5 μl of each amplification primer (0.3 pmol/μl), 2.5 μl of PCR-analyzed water, and 5 μl of DNA extract. Thermal cycling consisted of three repeated steps: a 3-minute hot-start polymerase activation-denaturation step at 95°C followed by 40 repeated cycles at 95°C for 10 seconds and then at 60°C for 30 seconds. Melting curves were obtained by increasing the temperature from 55 to 90°C with a plate reading every 0.2°C. In the PANIRINOX study, 28 different mutations on the *KRAS, BRAF* and *NRAS* genes actionable in mCRC management care are tested. The IntPlex assay is clinically validated^20,21^, and shows the highest sensitivity and specificity so far described^8^. Poisson law experiments demonstrated that this method could accurately detect down to one molecule per PCR reaction mixture^21^, with detection sensitivity as high as 1/100,000.

Intra- and inter-reproducibility experiments combining pre-analytic and analytic procedures demonstrated that the coefficient of variation for cirDNA concentration measurement is 24%^25^. IntPlex has been validated in two clinical studies^20,21^ is currently involved in 9 others, and has already enabled the testing of more than 2,000 individuals and 4,500 plama samples.

## 3. Statistics

Statistical analysis of pre-versus post-lockdown data was performed using the GraphPad Prism V6.01 software and survival analysis with STATA 16.0 software. Where appropriate, data were log transformed prior to statistical analysis. Continuous variables were compared using the Mann-Whitney test. Categorical variables were compared using the Pearson’s chi-square test. Median follow-up was calculated using the reverse Kaplan-Meier method. Overall survival (OS) was estimated using the Kaplan-Meier method, and compared using the Log-rank test. OS was defined as the time between the date of first metastatic diagnosis and the date of death from any cause. Hazard ratios (HR) are given with their 95% confidence interval (95% CI). Correlation analysis were performed using the spearman test. A probability of less than 0.05 was considered to be statistically significant; *p<0.05, **p<0.01;***p< 0.001; ****p< 0.0001.

## Supplementary figure legends

**Suppl. Figure 1:**
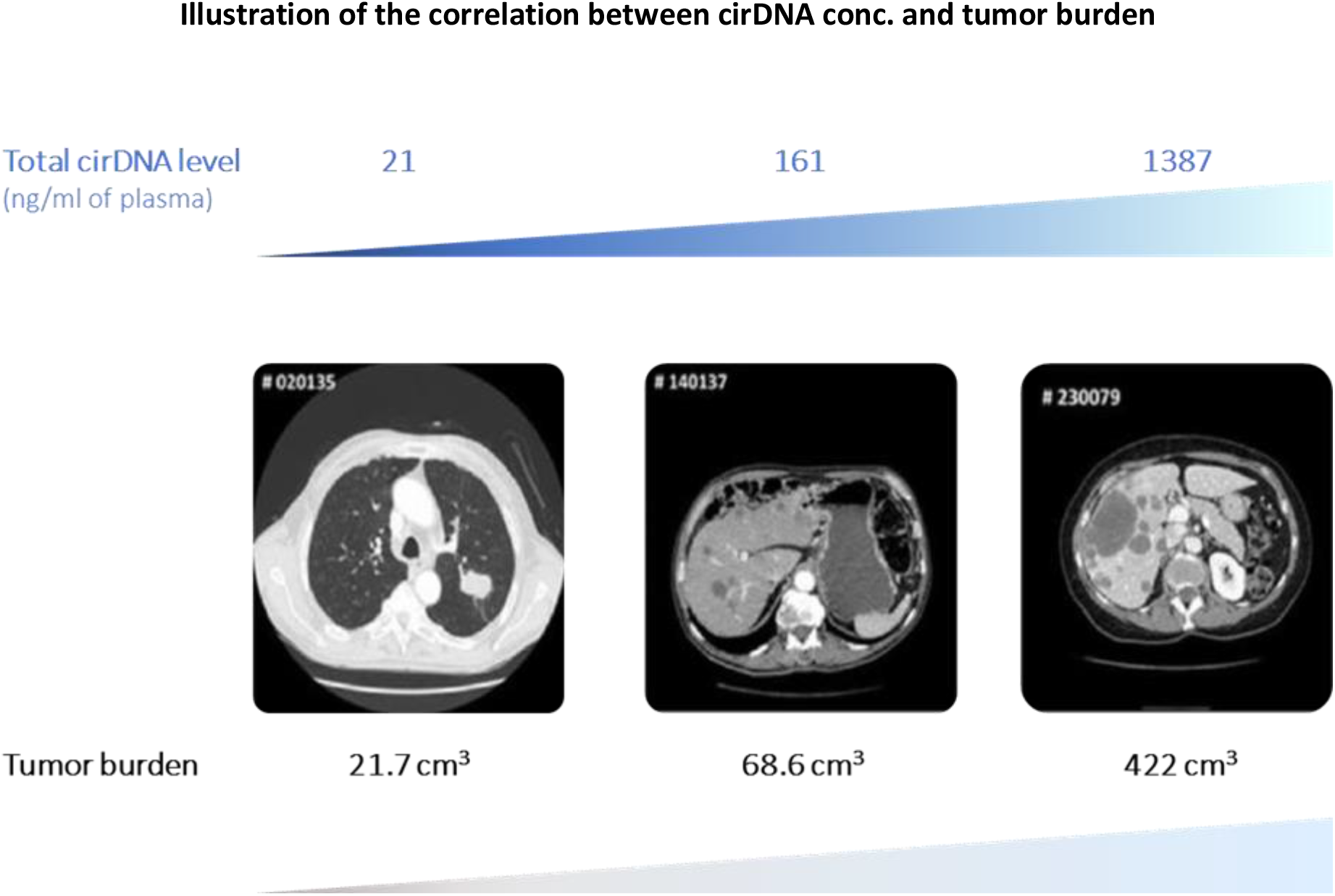
Illustration of the correlation between the tumor burden and total cirDNA level in three metachronous mCRC patients (one site) with increasing hepatic tumor mass as determined by MRI (adapted from our previous report: El Messaoudi et al, Clinical Cancer Research, 2013, ref #16).

Similar observation was reported by numerous other groups such as:

- Nygaard AD, Holdgaard PC, Spindler K-LG, Pallisgaard N, Jakobsen A. The correlation between cell-free DNA and tumour burden was estimated by PET/CT in patients with advanced NSCLC. Br J Cancer 2014;110:363–8
- Saluja, H.; Karapetis, C. S.; Pedersen, S. K.; Young, G. P.; Symonds, E. L. The Use of Circulating Tumor DNA for Prognosis of Gastrointestinal Cancers. Front. Oncol. 2018, 8, 275. https://doi.org/10.3389/fonc.2018.00275.
- Xu, X.; Yu, Y.; Shen, M.; Liu, M.; Wu, S.; Liang, L.; Huang, F.; Zhang, C.; Guo, W.; Liu, T. Role of Circulating Free DNA in Evaluating Clinical Tumor Burden and Predicting Survival in Chinese Metastatic Colorectal Cancer Patients. BMC Cancer 2020, 20 (1), 1006. https://doi.org/10.1186/s12885-020-07516-7.
- Hamfjord, J.; Guren, T. K.; Dajani, O.; Johansen, J. S.; Glimelius, B.; Sorbye, H.; Pfeiffer, P.; Lingjærde, O. C.; Tveit, K. M.; Kure, E. H.; Pallisgaard, N.; Spindler, K.-L. G. Total Circulating Cell-Free DNA as a Prognostic Biomarker in Metastatic Colorectal Cancer before First-Line Oxaliplatin-Based Chemotherapy. Annals of Oncology 2019, 30 (7), 1088–1095. https://doi.org/10.1093/annonc/mdz139.

**Suppl. Figure 2:**
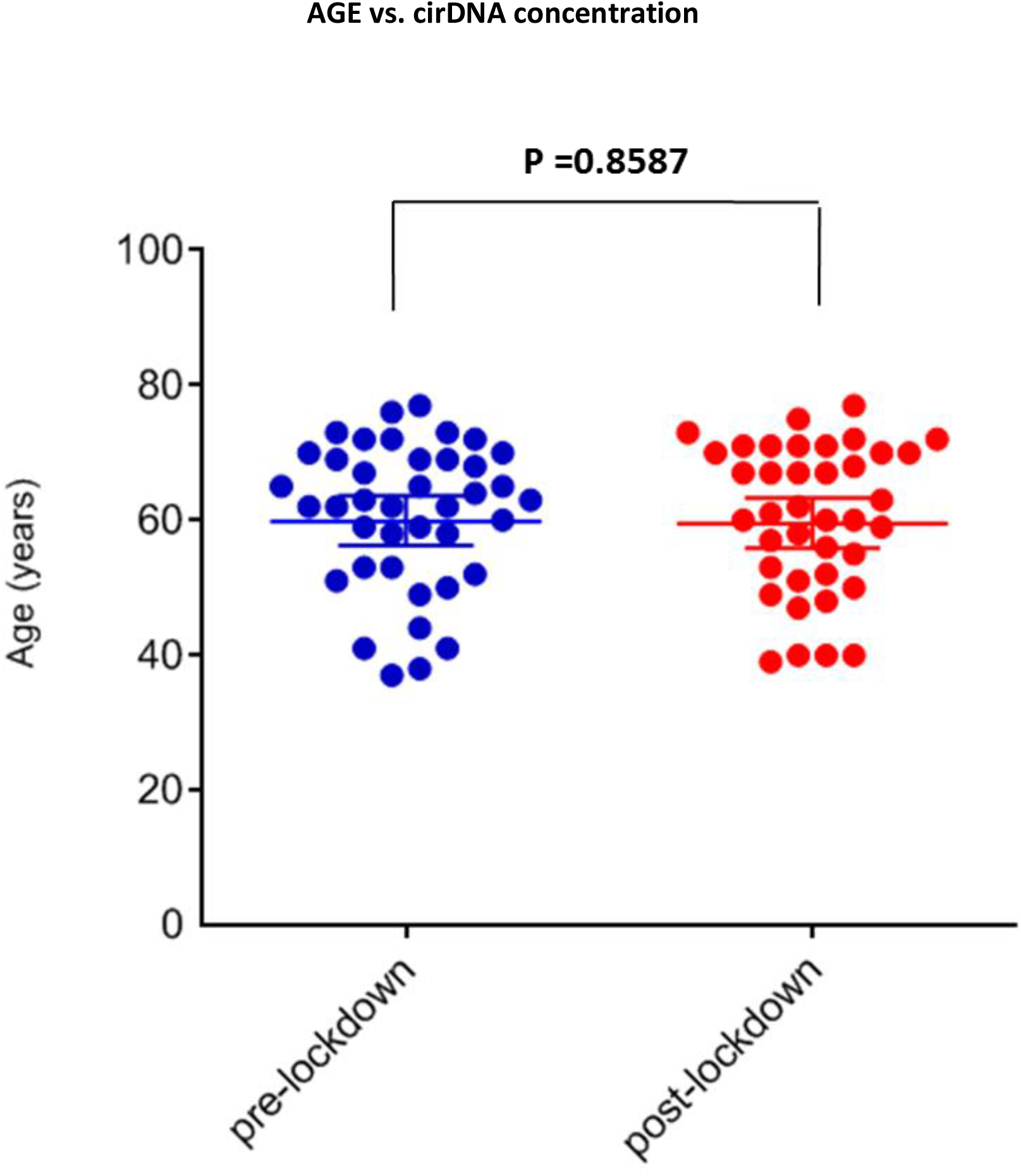
Comparison of the age of the newly diagnosed mCRC patients from the pre- and post-lockdown study cohorts (N=80). Group median is represented by a horizontal bold bar. Mann-Whitney U test was performed to compare distribution in pre- and post-lockdown patients. Each dot (blue, pre-lockdown; red, post-lockdown) represents the values of a single patient.

**Suppl. Figure 3:**
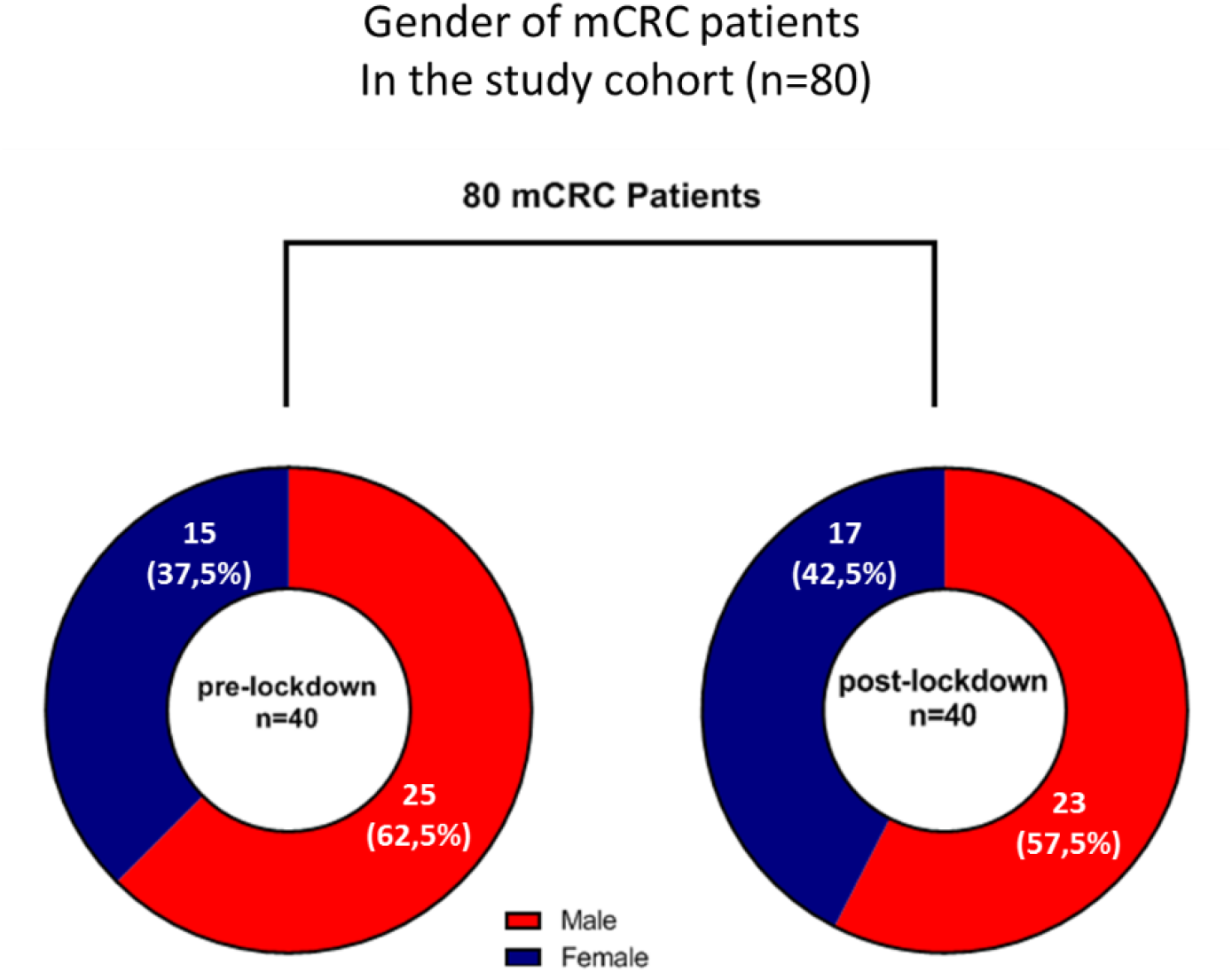
Comparison of the gender of the newly diagnosed mCRC patients from the pre- and post-lockdown study cohorts (N=80).

**Suppl. Figure 4:**
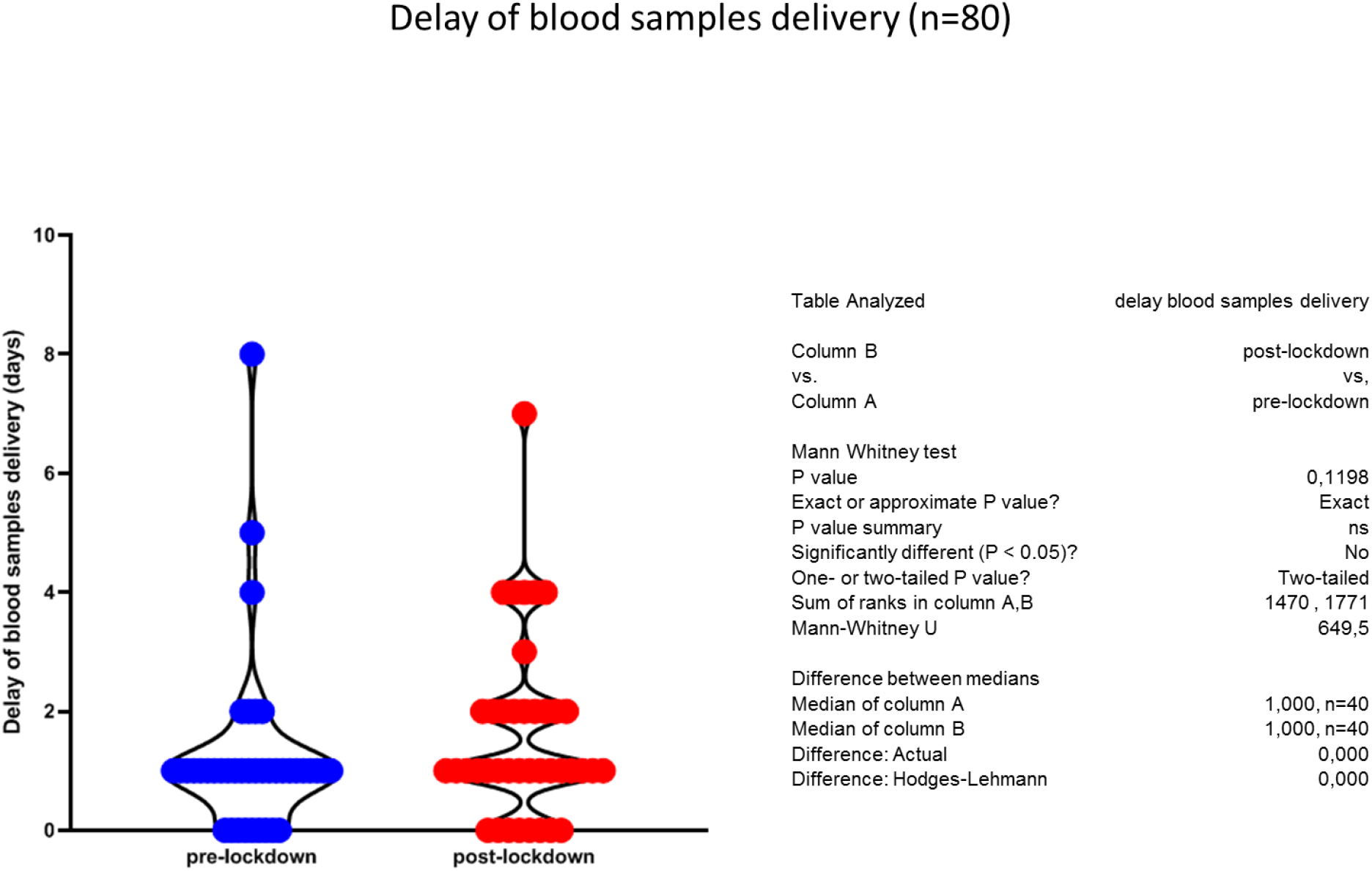
Comparison of the delivery delay of blood samples from the newly diagnosed mCRC patients from the pre- and post-lockdown study cohorts. Mann-Whitney U test was performed to compare distribution in pre- and post-lockdown patients. Each dot (blue, pre-lockdown; red, post-lockdown) represents the values of a single patient.

**Suppl. Figure 5:**
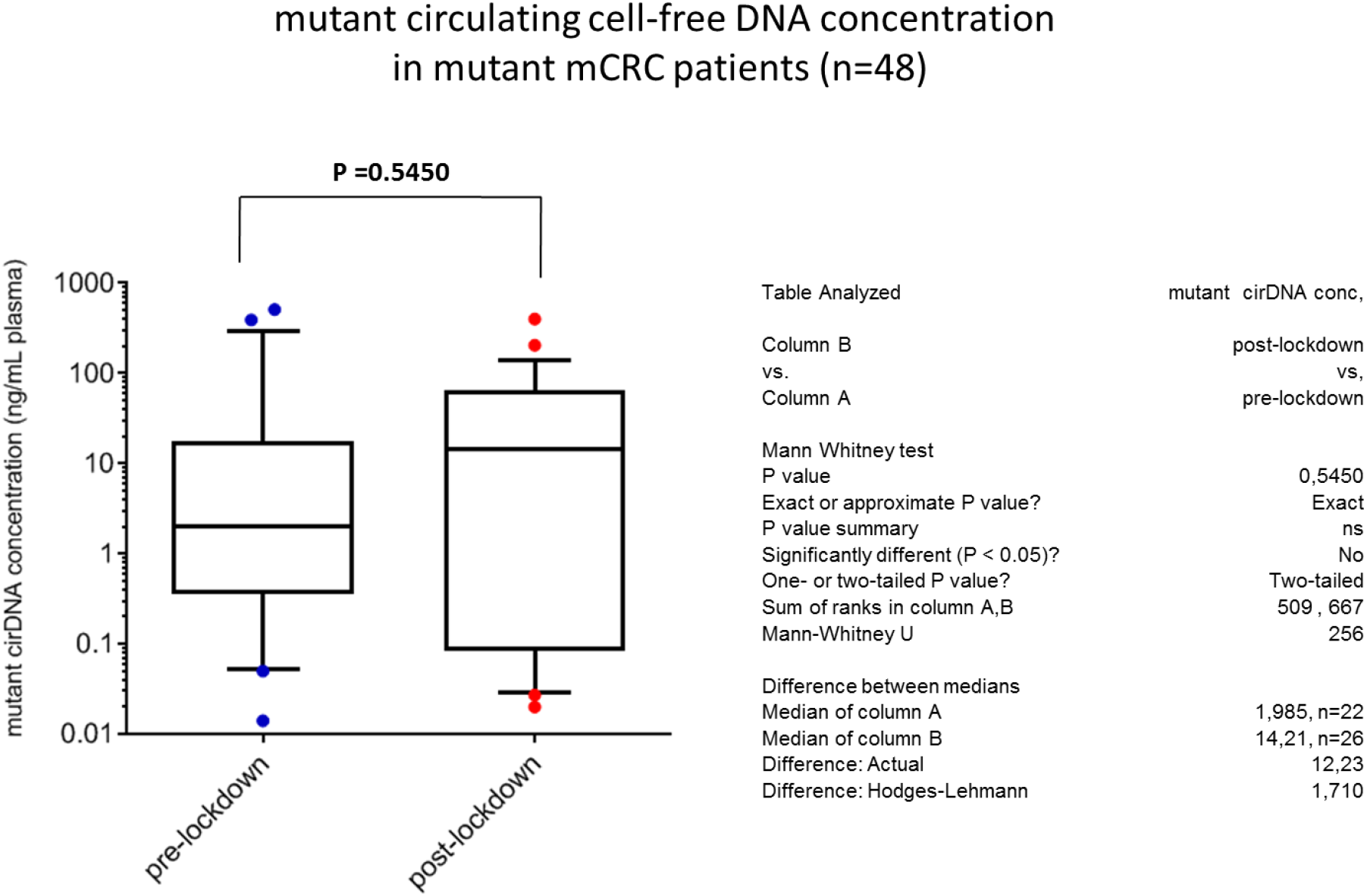
Comparison of the mutant cirDNA concentration in the newly mutant diagnosed mCRC patients from pre-and post-lockdown study cohorts (N=48). Group median is represented by a horizontal bold bar. Mann-Whitney U test was performed to compare distribution in pre- and post-lockdown patients. Each dot (blue, pre-lockdown; red, post-lockdown) represents the values of a single patient.

**Suppl. Figure 6:**
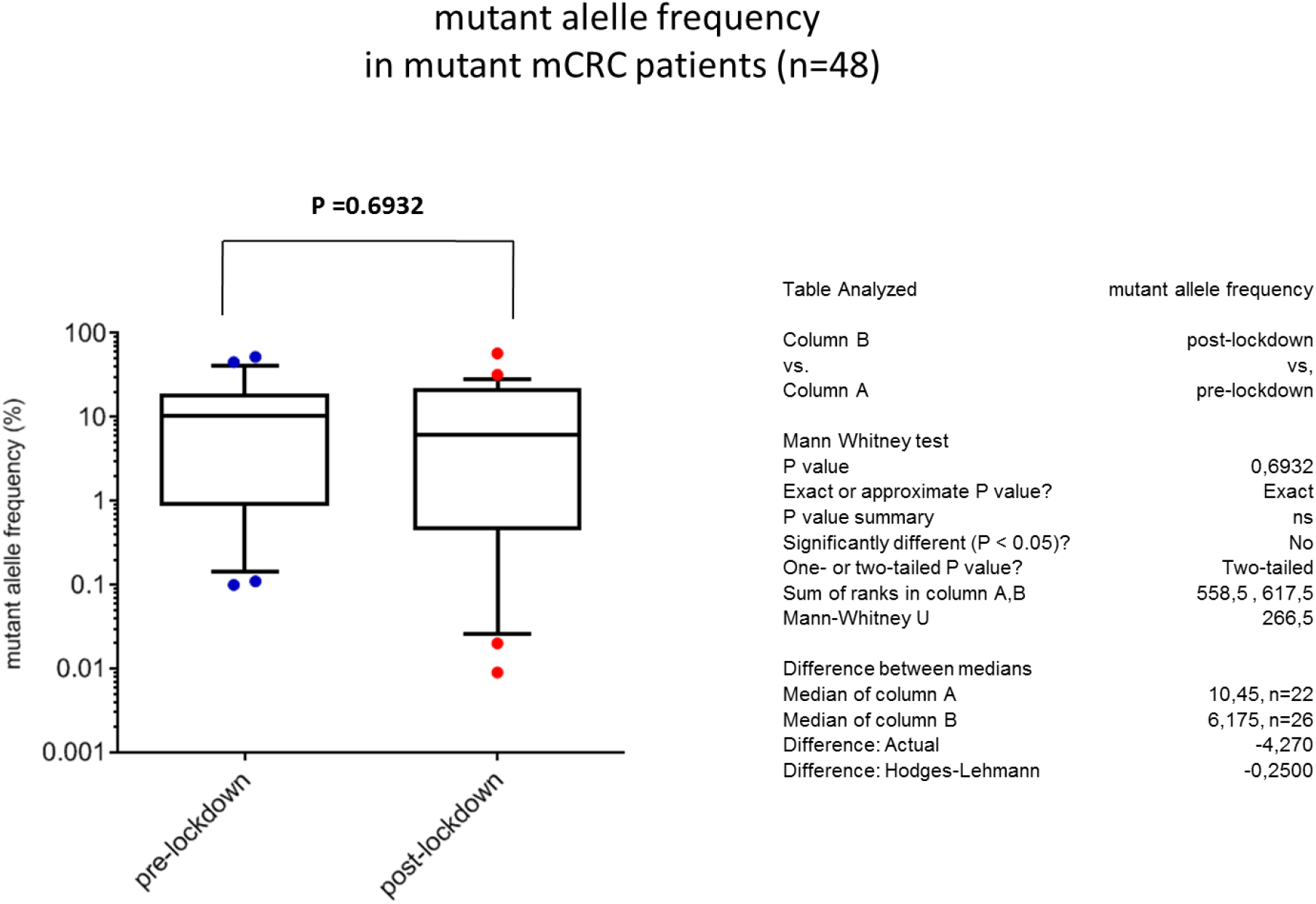
Comparison of the mutant allele frequency in the newly diagnosed mutant mCRC patients from the pre- and post-lockdown study cohort (N=48). Group median is represented by a horizontal bold bar. Mann-Whitney U test was performed to compare distribution in pre- and post-lockdown patients. Each dot (blue, pre-lockdown; red, post-lockdown) represents the values of a single patient.

**Suppl. Figure 7:**
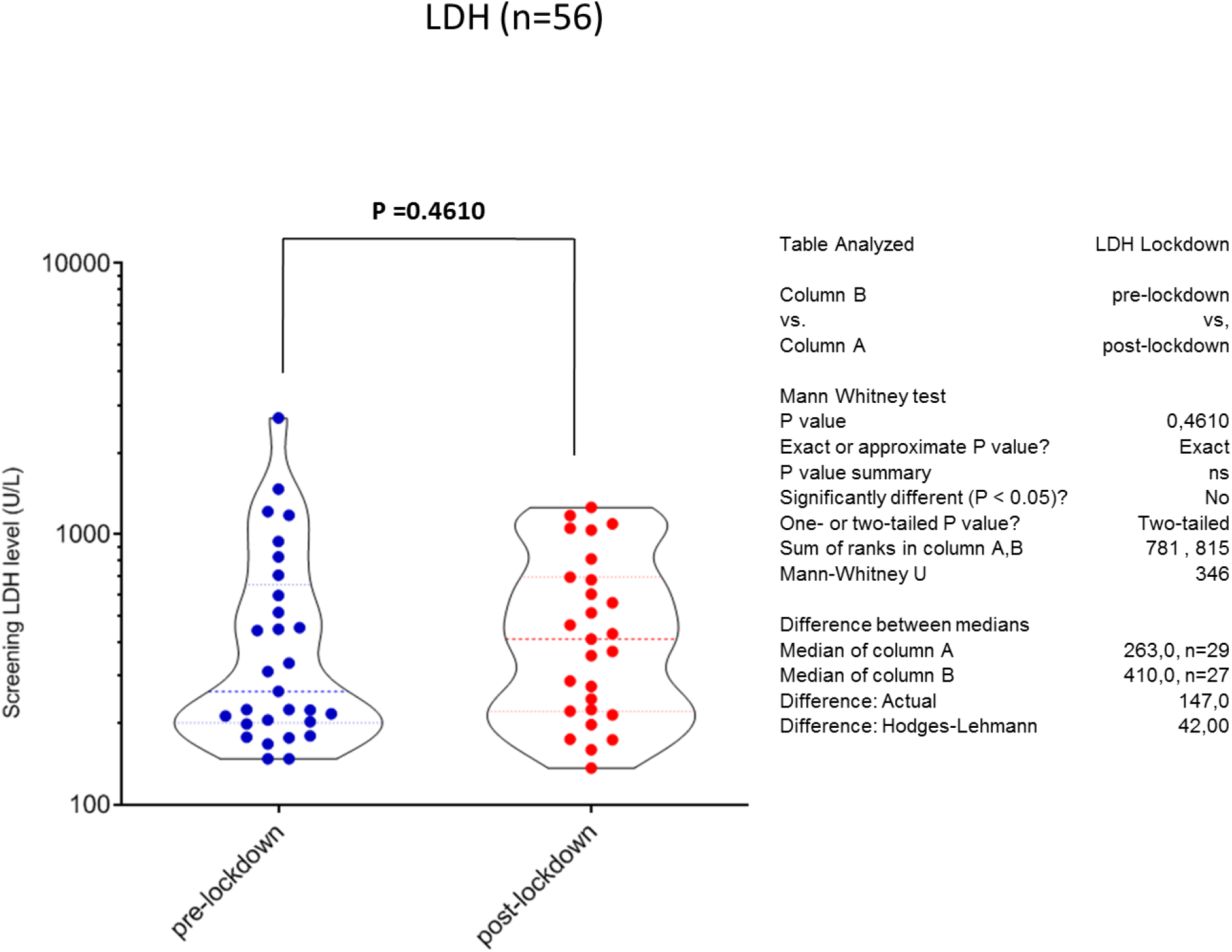
Comparison of the Lactate dehydrogenase (LDH) of the newly diagnosed mCRC patients from the pre- and post-lockdown study cohorts (N=56). Group median is represented by a horizontal bold dotted line; violin representation. Mann-Whitney U test was performed to compare distribution in pre- and post-lockdown patients. Each dot (blue, pre-lockdown; red, post-lockdown) represents the values of a single patient.

**Suppl. Figure 8:**
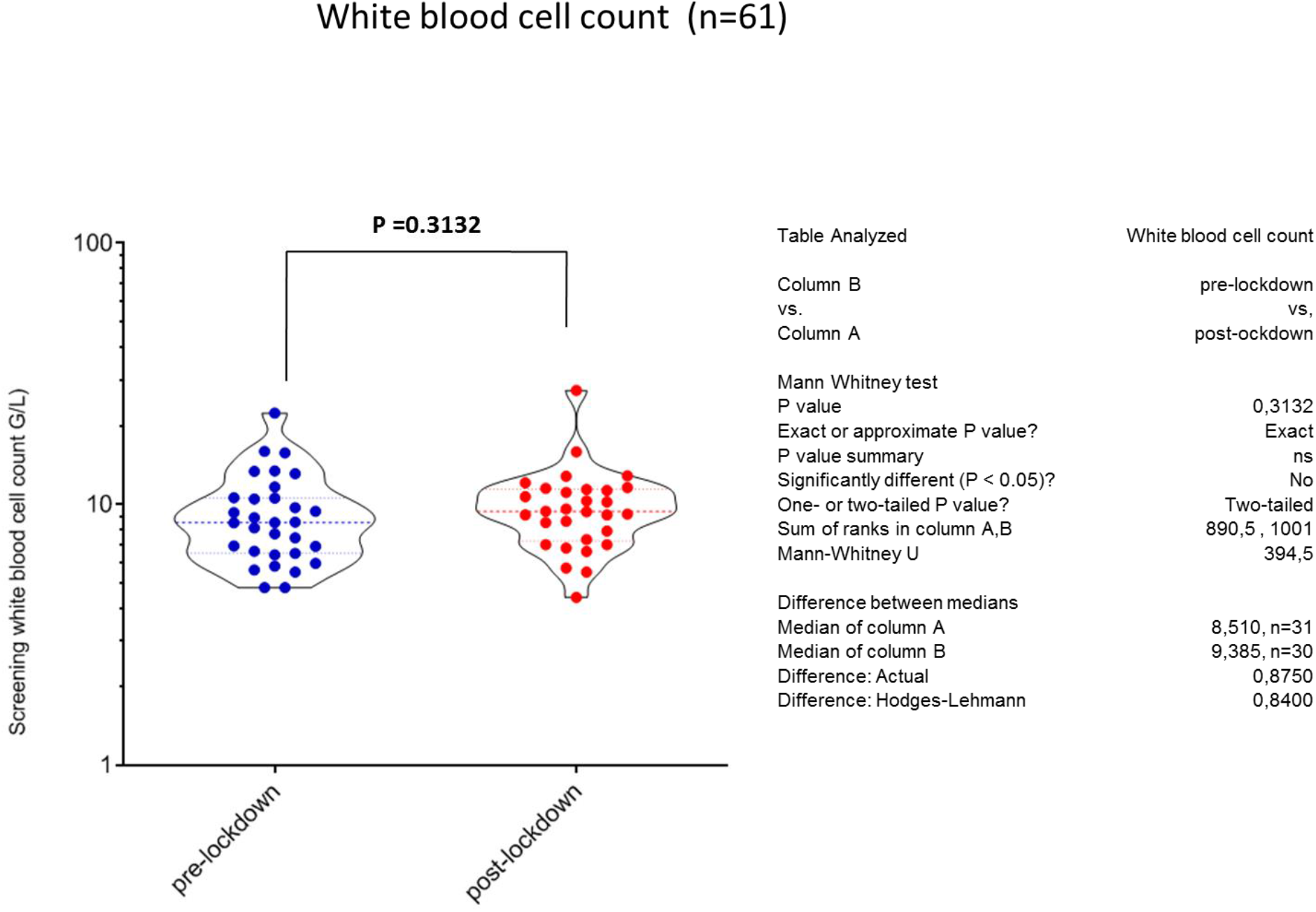
Comparison of the white blood cell count in the newly diagnosed mCRC patients from the pre- and post-lockdown study cohorts (N=61). Group median is represented by a horizontal bold dotted line; violin representation. Mann-Whitney U test was performed to compare distribution in pre- and post-lockdown patients. Each dot (blue, pre-lockdown; red, post-lockdown) represents the values of a single patient.

**Suppl. Figure 9:**
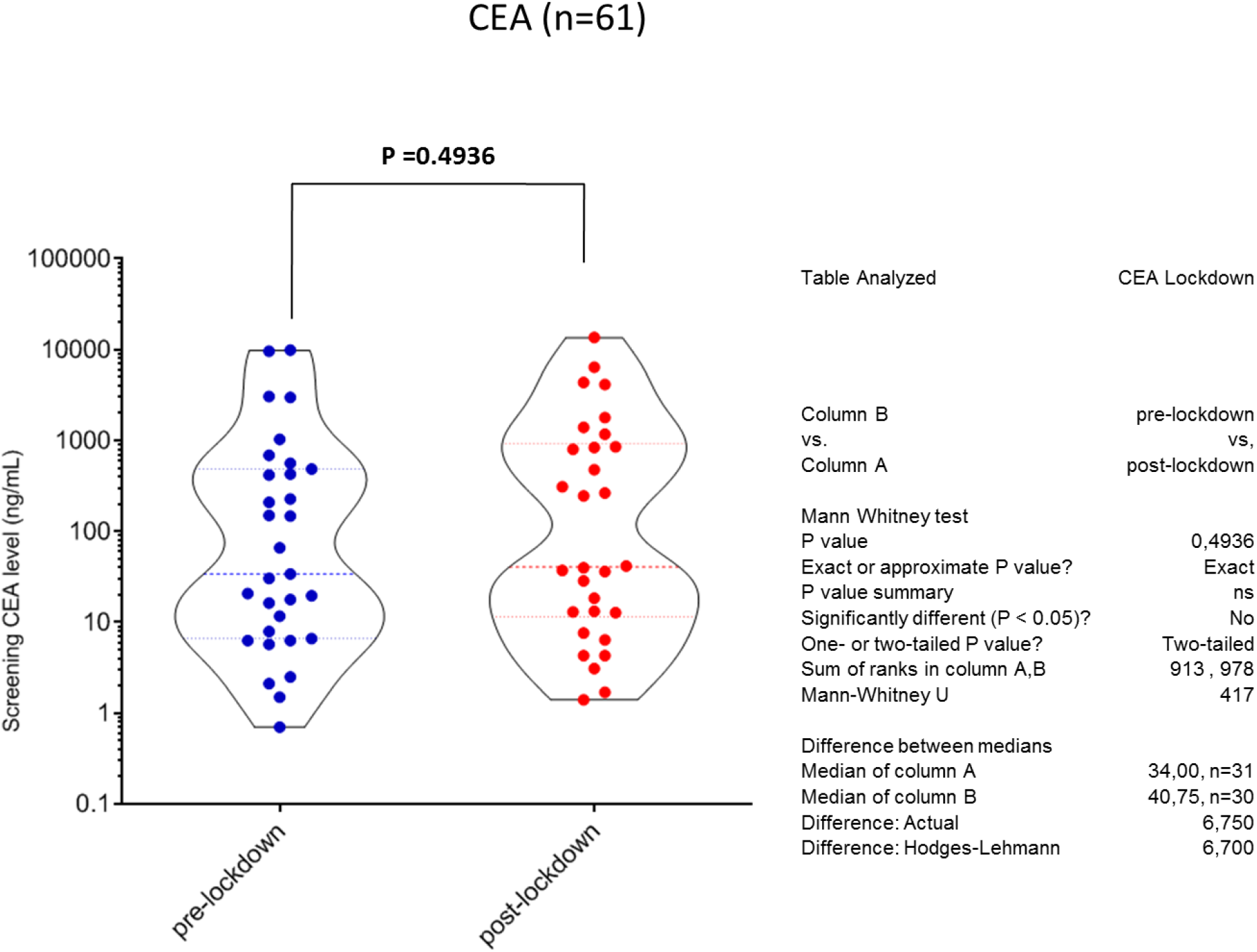
Comparison of the carcinoembryonic antigen (CEA) in the newly diagnosed mCRC patients from the pre- and post-lockdown study cohorts (N=61). Group median is represented by a horizontal bold dotted line; violin representation. Mann-Whitney U test was performed to compare distribution in pre- and post-lockdown patients. Each dot (blue, pre-lockdown; red, post-lockdown) represents the values of a single patient.

**Suppl. Figure 10:**
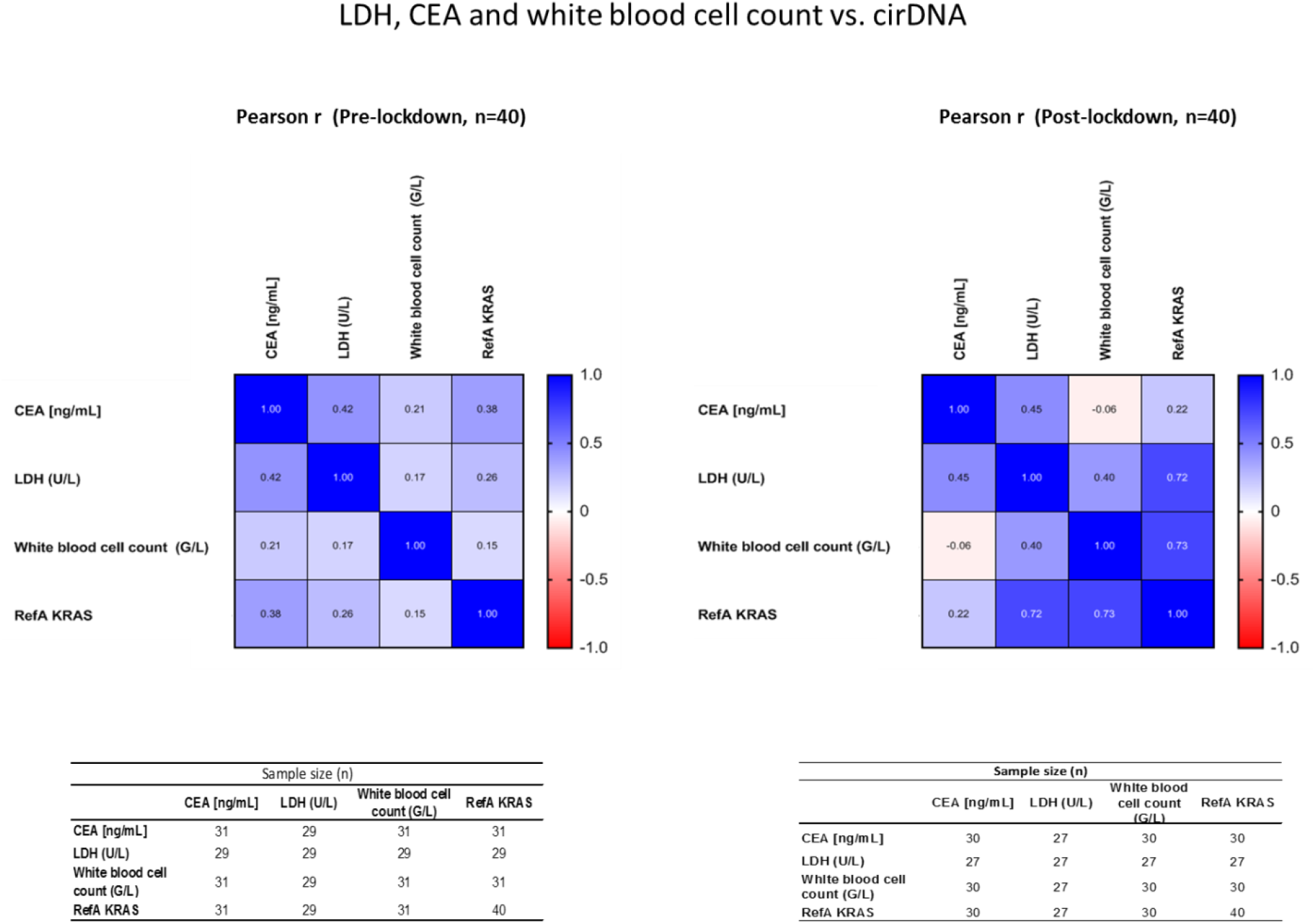
Pearson r correlation analysis of the cirDNA, LDH, white blood cell count, and CEA levels in the newly diagnosed mCRC patients from the pre- and post-lockdown study cohorts.

**Suppl. Figure 11:**
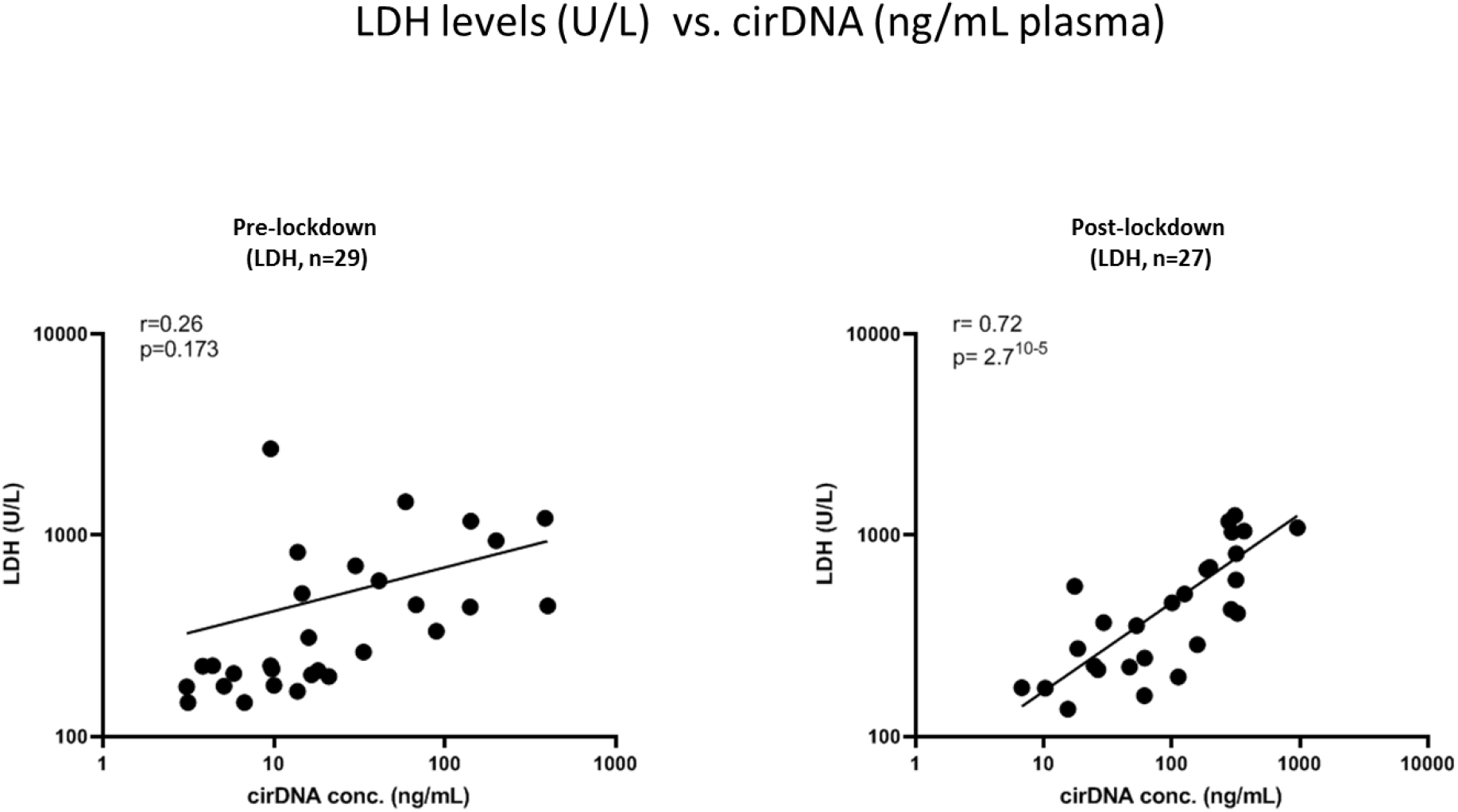
Scatter plots showing the correlation between LDH and cirDNA concentrations in pre and post-lockdown cohorts. Each dot represents the values of a single patient; slanting lines are the regression lines as performed by the Spearman test.

**Suppl. Figure 12:**
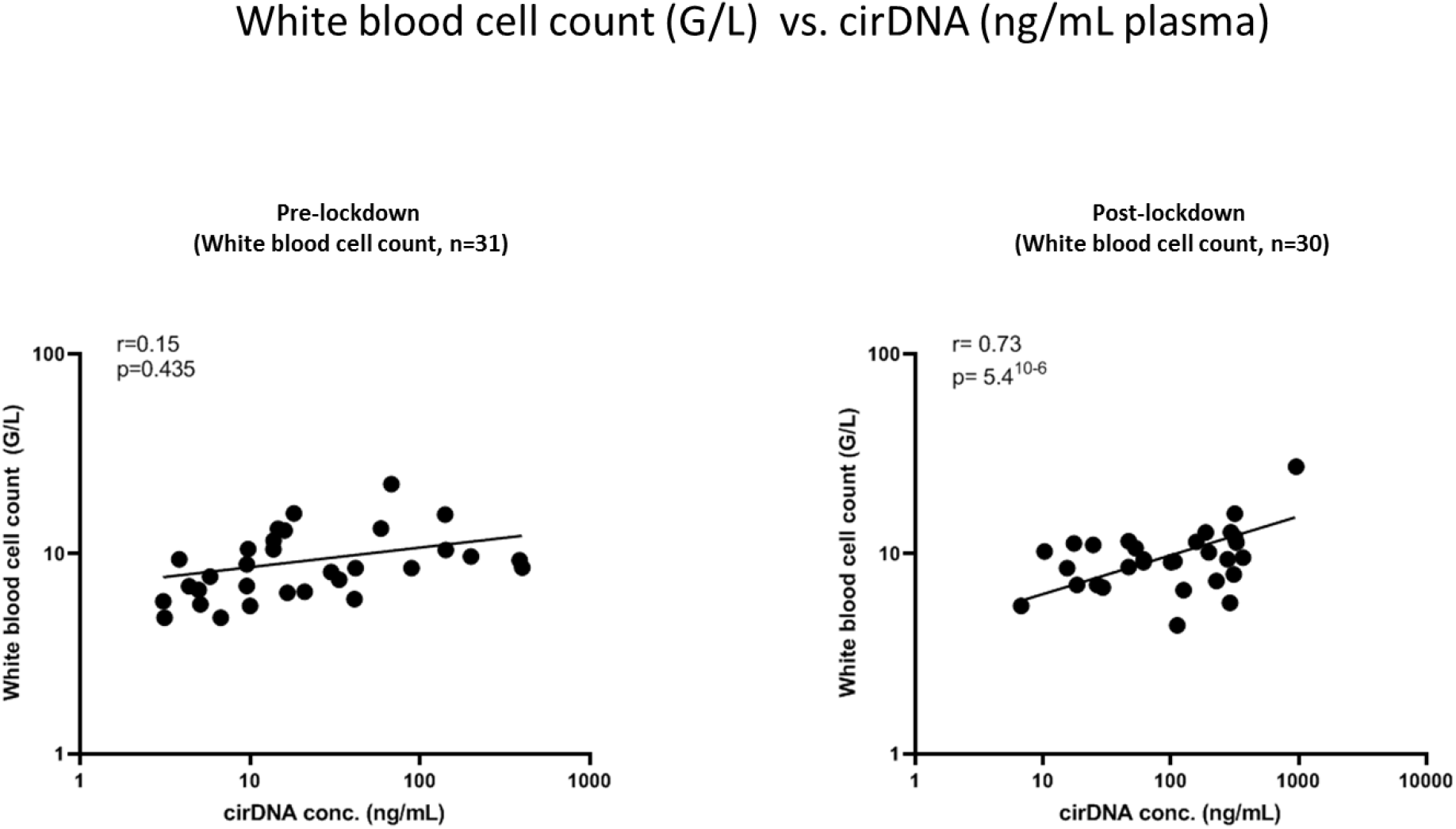
Scatter plots showing the correlation between white blood cell count and cirDNA concentration in pre and post-lockdown cohorts. Each dot represents the values of a single patient; slanting lines are the regression lines as performed by the Spearman test.

**Suppl. Figure 13:**
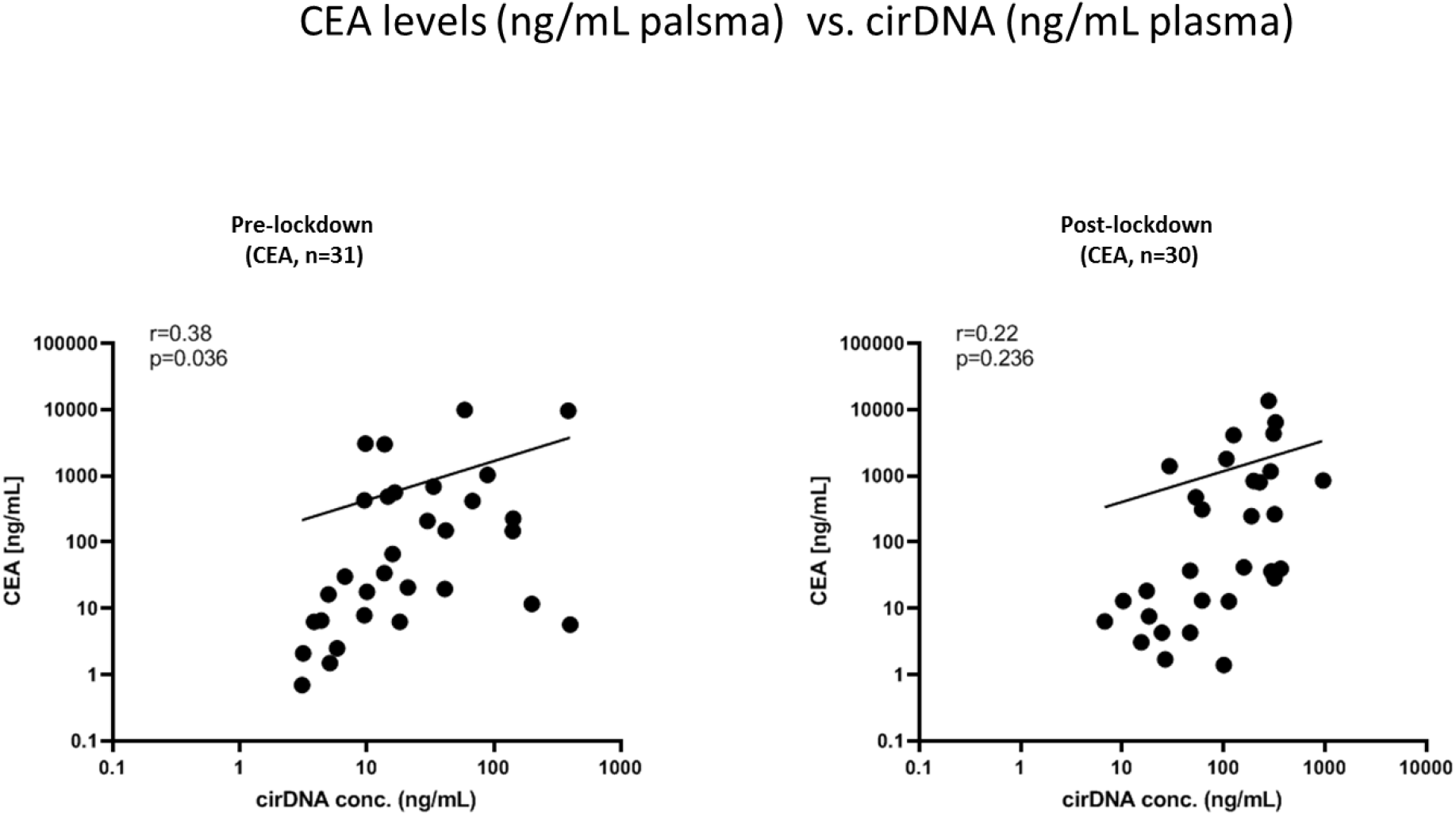
Scatter plots showing the correlation between CEA and cirDNA concentrations in pre and post-lockdown cohorts. Each dot represents the values of a single patient; slanting lines are the regression lines as performed by the Spearman test.

**Suppl. Figure 14:**
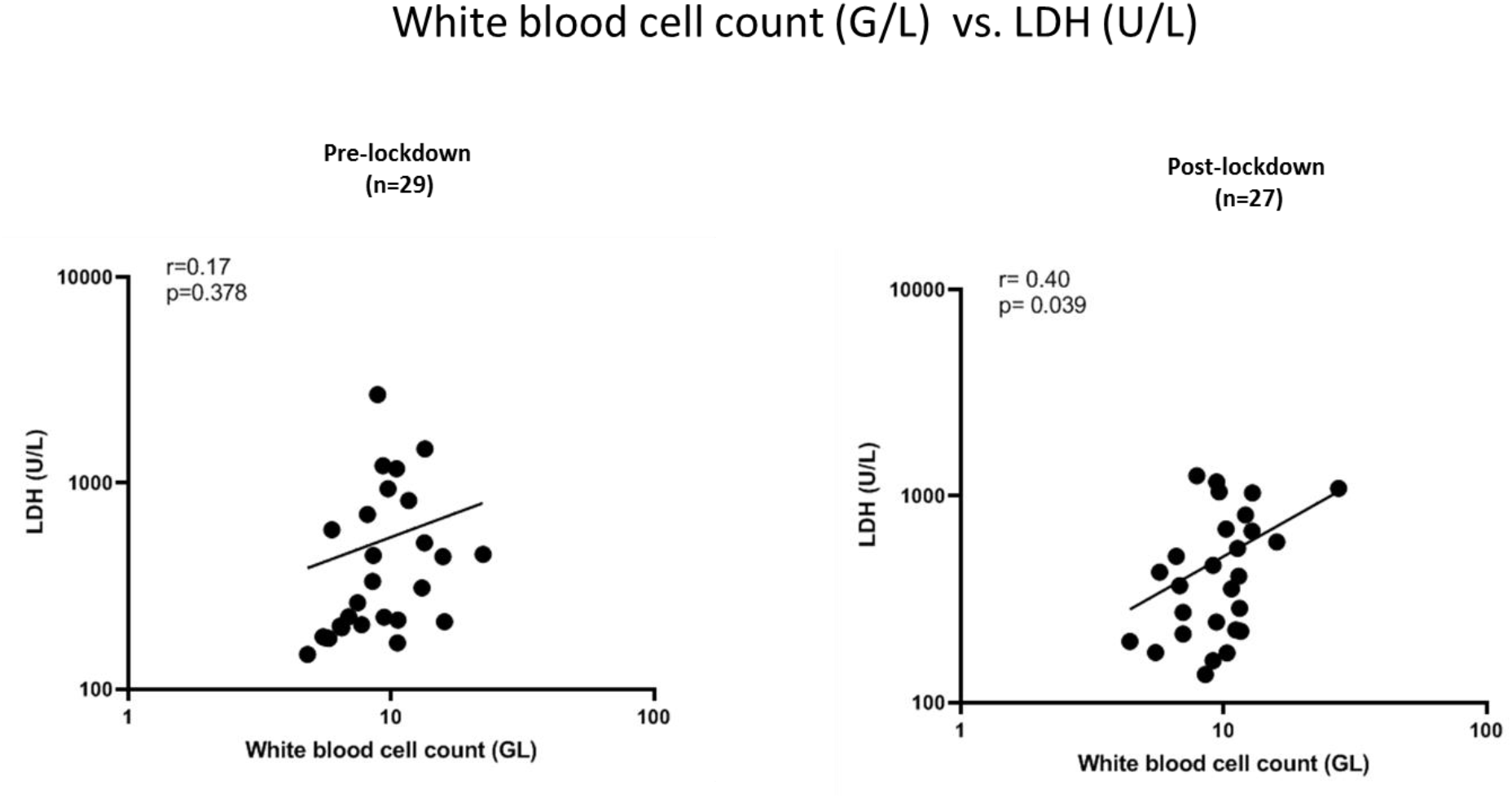
Scatter plots showing the correlation between white blood cell count and LDH concentration in pre and post-lockdown cohorts. Each dot represents the values of a single patient; slanting lines are the regression lines as performed by the Spearman test.

**Supplementary Table 1.**
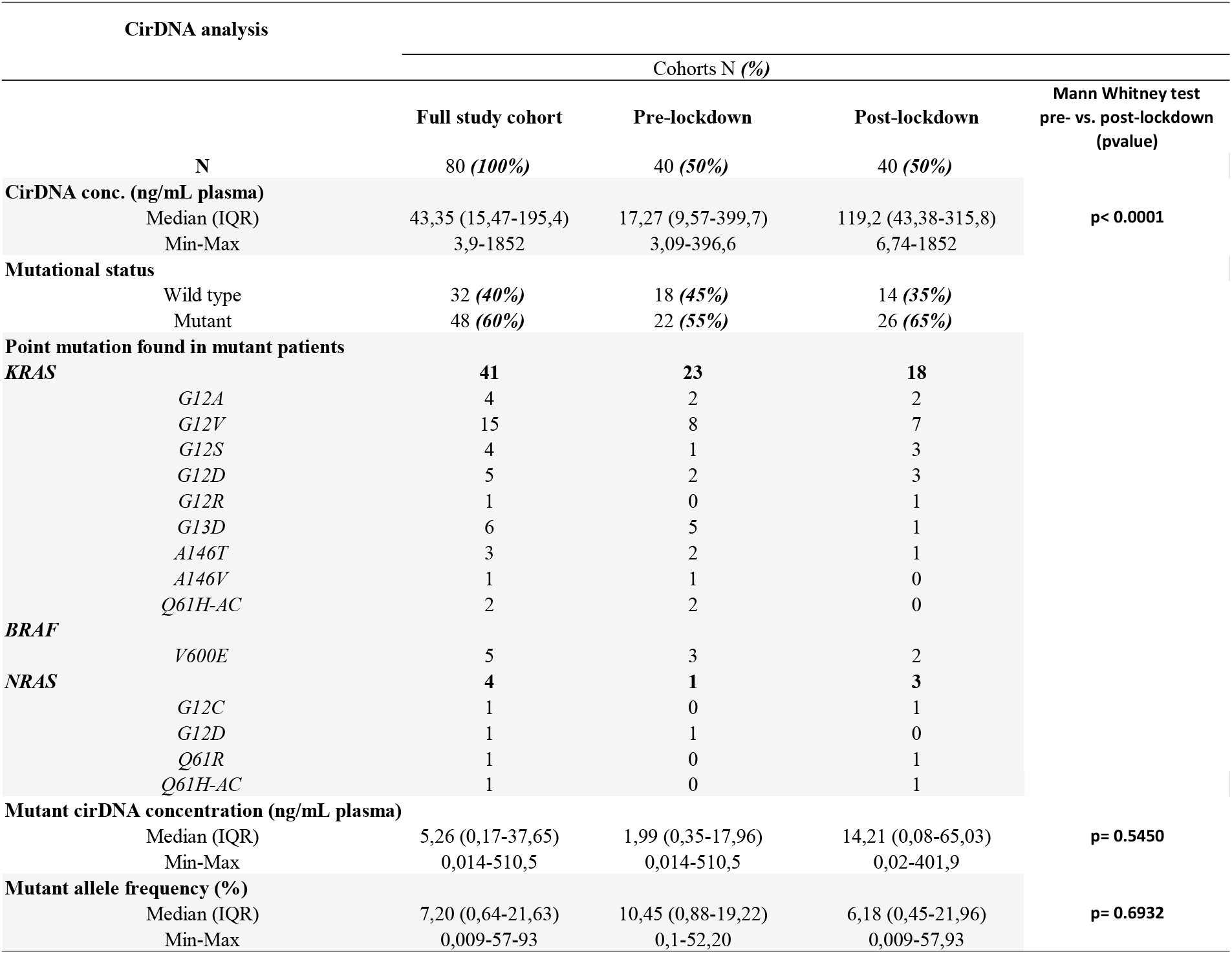

**Supplementary Table 2.**
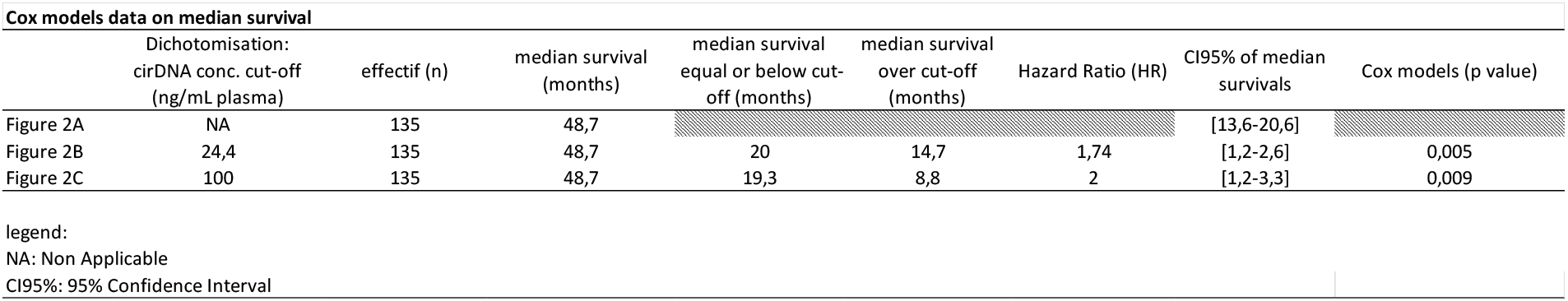

